# Observed serial intervals of SARS-CoV-2 for the Omicron and Delta variants in Belgium based on contact tracing data, 19 November to 31 December 2021

**DOI:** 10.1101/2022.01.28.22269756

**Authors:** Cécile Kremer, Toon Braeye, Kristiaan Proesmans, Emmanuel André, Andrea Torneri, Niel Hens

**Author notes:** These authors contributed equally (Cécile Kremer, Toon Braeye).

## Abstract

The SARS-CoV-2 Omicron BA.1 variant is rapidly spreading worldwide, possibly outcompeting the Delta strain. We investigated the empirical serial interval for both variants using contact tracing data. Overall, we observed a shorter serial interval for Omicron compared to Delta, suggesting faster transmission. Furthermore, results indicate a relation between the empirical serial interval and the vaccination status for both the Omicron and the Delta variant. Consequently, with the progression of the vaccination campaign, the reasons for and extent of dominance of Omicron over Delta may need further assessment.

## Background

The WHO designated the SARS-CoV-2 Omicron BA.1 variant (B.1.1.529) as a variant of concern (VoC) on 26 November 2021 [1]. Omicron shows a fast epidemic growth and has taken over as dominant VoC from the previously dominant Delta variant (B.1.617.2) worldwide. In Belgium, the Omicron variant was the dominant circulating strain during the period from 27 December 2021 to 9 January 2022, identified in 88.5% of the sequenced samples [2]. It has been found that the Omicron variant is more efficient at evading immunity, acquired from previous infection or vaccination [3,4], compared to the Delta variant. Another epidemiological characteristic that may contribute to the rapid spread of Omicron is increased transmissibility, possibly due to an increase in the reproduction number or a shortened serial interval, i.e. the time between symptom onset in an infector and infectee [5]. In this study, we estimate the mean and the standard deviation of the serial interval for both the Omicron and Delta variants using contact tracing data collected in Belgium and assess whether these variants are associated with a different observed serial interval. To gain more insights on the possible impact of vaccination, we also compare the observed serial intervals for different combinations of vaccination status in transmission pairs.

### Data

Belgium has a contact tracing system in place, where COVID-19 confirmed cases are asked about their contacts during two days prior to symptom onset until 10 days after. Variant detection was done using genotype sequencing. If a variant was found in a transmission chain, all cases belonging to that chain were assumed to be infected by that variant. We collected transmission pairs that could be linked either to Omicron or Delta infections, where the infector reported first symptoms between 19 November and 31 December 2021. During this period Omicron started to spread in Belgium and took over dominance from the Delta variant [6]. The same non-pharmaceutical interventions were in place throughout the considered time window, with a fairly low stringency index (around 48) compared to neighboring countries.We assumed that the first confirmed case in a reconstructed transmission pair (i.e. the ‘index’ case) was the infector, and hence the contact was the infectee. Transmission pairs for which symptom onset was not available for either the infector or the infectee were excluded, as well as pairs for which the observed serial interval was less than -5 days or more than 15 days in order to ensure biologically plausible serial intervals [7,8]. Vaccination status was assigned to both cases in a transmission pair as (i) unvaccinated (including partially vaccinated), (ii)vaccinated (i.e. completed vaccination cycle), and (iii) vaccinated plus booster. Household status was assigned based on information collected during the interview with the index case or provided by the healthcare worker. A more detailed description of the data and analysis is provided in Supplement S1.

### Overall serial interval

Of the 2495 included transmission pairs, 86.61% were linked to transmission of the Omicron variant (Figure S1). In what follows we will report the mean and standard deviation (SD) of the observed serial intervals, the median was 3 days for all stratifications. All reported *p*-values are based on a Mann-Whitney U test. Tables S1 and S2 show the proportion of transmission pairs by household and vaccination status.

We identified 2161 Omicron and 334 Delta transmission pairs during the period from 19 November to 31 December 2021. The empirical serial interval distribution for Omicron had a mean of 2.75 days (SD 2.53 days), compared to 3.00 days (SD 2.48 days) for Delta (*p* = .019; Figures 1A-B). Table 1 shows the parameter estimates of the normal distribution fit to both empirical serial interval distributions (Figure 1C).

**Table 1.** Estimated parameters of a normal distribution for the serial interval of Omicron and Delta variants, by different stratifications. There were no Delta transmission pairs in which both cases had received a booster vaccine. Reported estimates are the posterior median and 95% credible interval (CrI).

| Variant | Stratification | Mean (95%CrI) | Standard deviation (95%CrI) |
| --- | --- | --- | --- |
| Omicron | / | 2.75 (2.65 – 2.86) | 2.54 (2.46 – 2.61) |
| Delta | / | 3.00 (2.73 – 3.26) | 2.49 (2.31 – 2.69) |
| Omicron | Within-household | 2.80 (2.67 – 2.93) | 2.60 (2.50 – 2.70) |
| Delta | Within-household | 3.04 (2.75 – 3.33) | 2.43 (2.24 – 2.65) |
| Omicron | Between-household | 2.72 (2.53 – 2.90) | 2.44 (2.31 – 2.57) |
| Delta | Between-household | 2.78 (2.00 – 3.56) | 2.78 (2.30 – 3.45) |
| Omicron | Both unvaccinated | 2.69 (2.40 – 2.98) | 2.75 (2.55 – 2.96) |
| Delta | Both unvaccinated | 2.54 (1.96 – 3.12) | 2.83 (1.93 – 2.77) |
| Omicron | Both vaccinated | 2.63 (2.46 – 2.81) | 2.45 (2.33 – 2.58) |
| Delta | Both vaccinated | 3.38 (2.89 – 3.88) | 2.47 (2.16 – 2.86) |
| Omicron | Both vaccinated + booster | 3.34 (2.58 – 4.10) | 2.59 (2.13 – 3.24) |
| Delta | Both vaccinated + booster | Not available | Not available |

**Figure 1.**
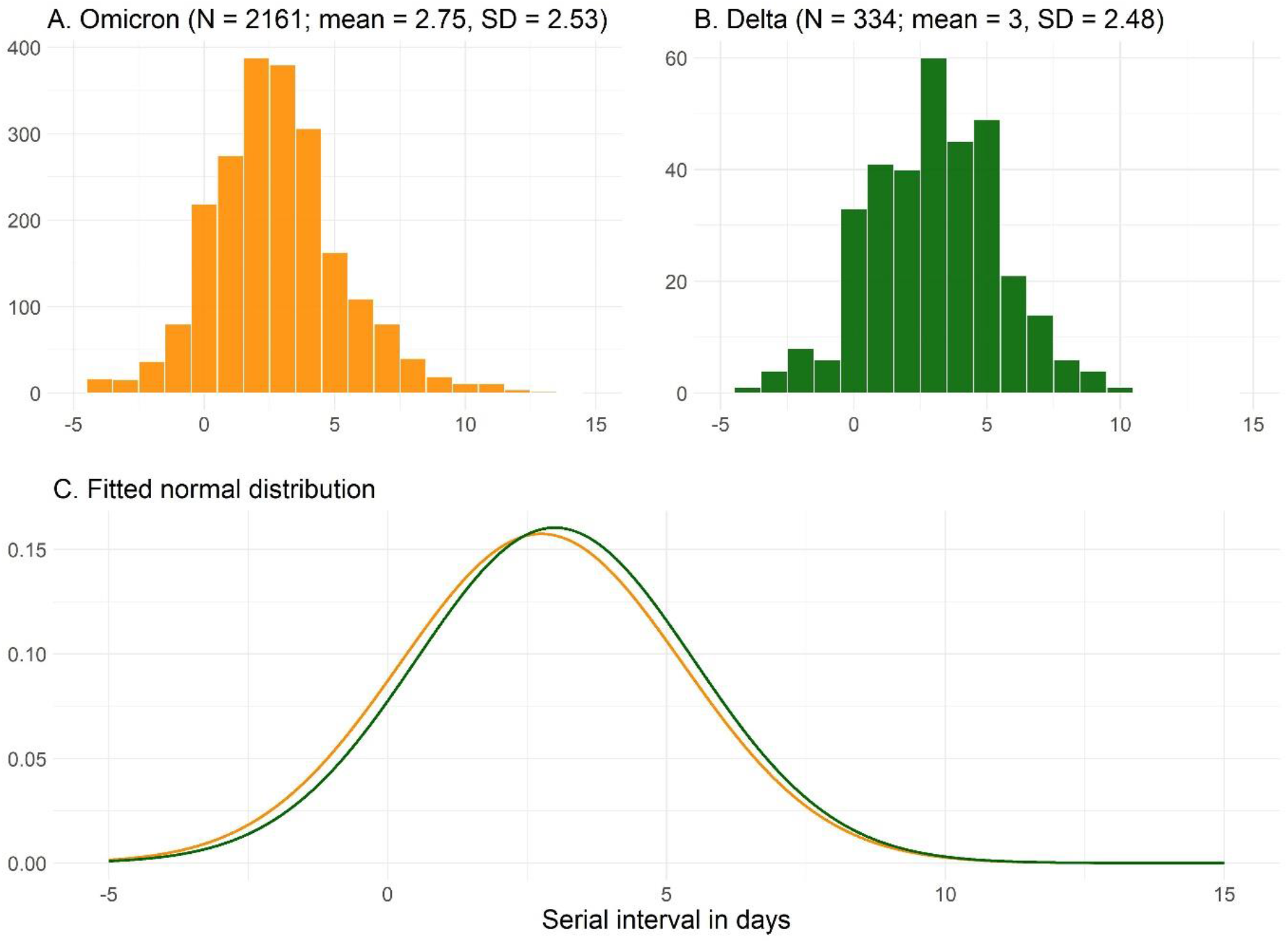
Empirical (A-B) and fitted normal (C) distribution of the serial intervals for Omicron and Delta variants. Onset date infector from 19 November until 31 December 2021.

### Serial interval stratified by household status

There was a significant difference between the mean empirical serial interval of both variants within households, but not between households (*p* = .034 for within-household, and *p* = .686 for between-household transmission pairs; Figures 2A-D). Figure 2E represents the fit of a normal distribution to the empirical serial intervals for both variants, and in Table 1 we report the parameters of these fitted distributions.

**Figure 2.**
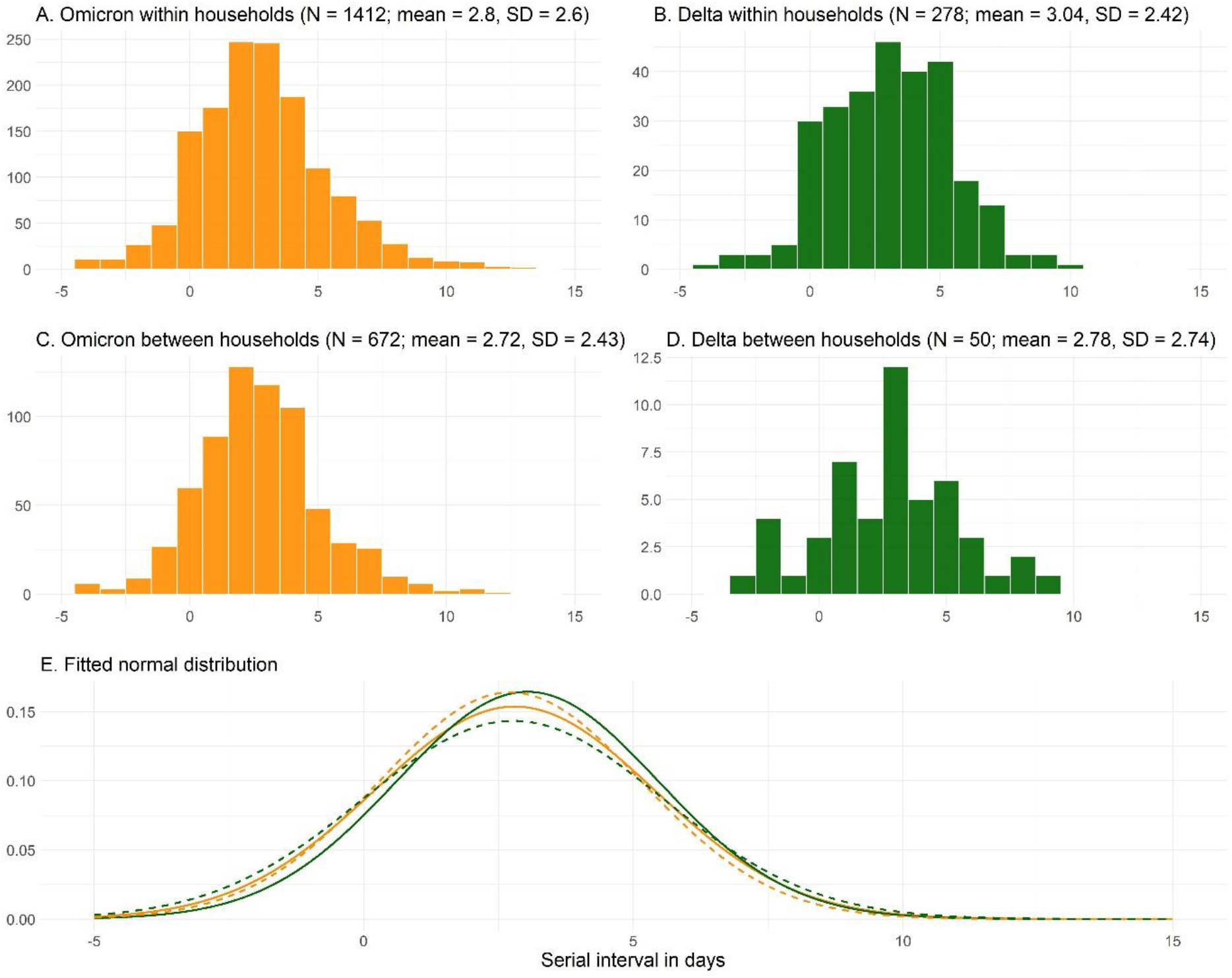
Empirical (A-D) and fitted normal (E; solid = within-household, dashed = between-household) distribution of the serial intervals for Omicron and Delta variant, by household status (excluding 83 pairs for which no information on household status was available). Onset date infector from 19 November until 31 December 2021.

### Serial interval stratified by vaccination status

No difference in empirical serial intervals was found for pairs where both cases were unvaccinated or only partially vaccinated (2.69 vs 2.54 days, *p* = .931; Figure 3A-B). For transmission pairs in which both cases were vaccinated (without booster), the empirical serial interval for Omicron was significantly shorter compared to Delta (2.63 vs 3.38 days, *p* = .004; Figure 3C-D). The mean empirical serial interval for Omicron was longer for pairs that received a booster vaccine compared to pairs that were vaccinated with only two doses (3.34 vs 2.63 days, *p* = .065; Figure 3C-E). The mean empirical serial interval was significantly longer for the Delta variant in transmission pairs in which both cases completed the vaccination cycle, compared to those where both cases were unvaccinated or only partially vaccinated (3.38 vs. 2.54 days, *p* = .045; Figure 3B-D).

**Figure 3.**
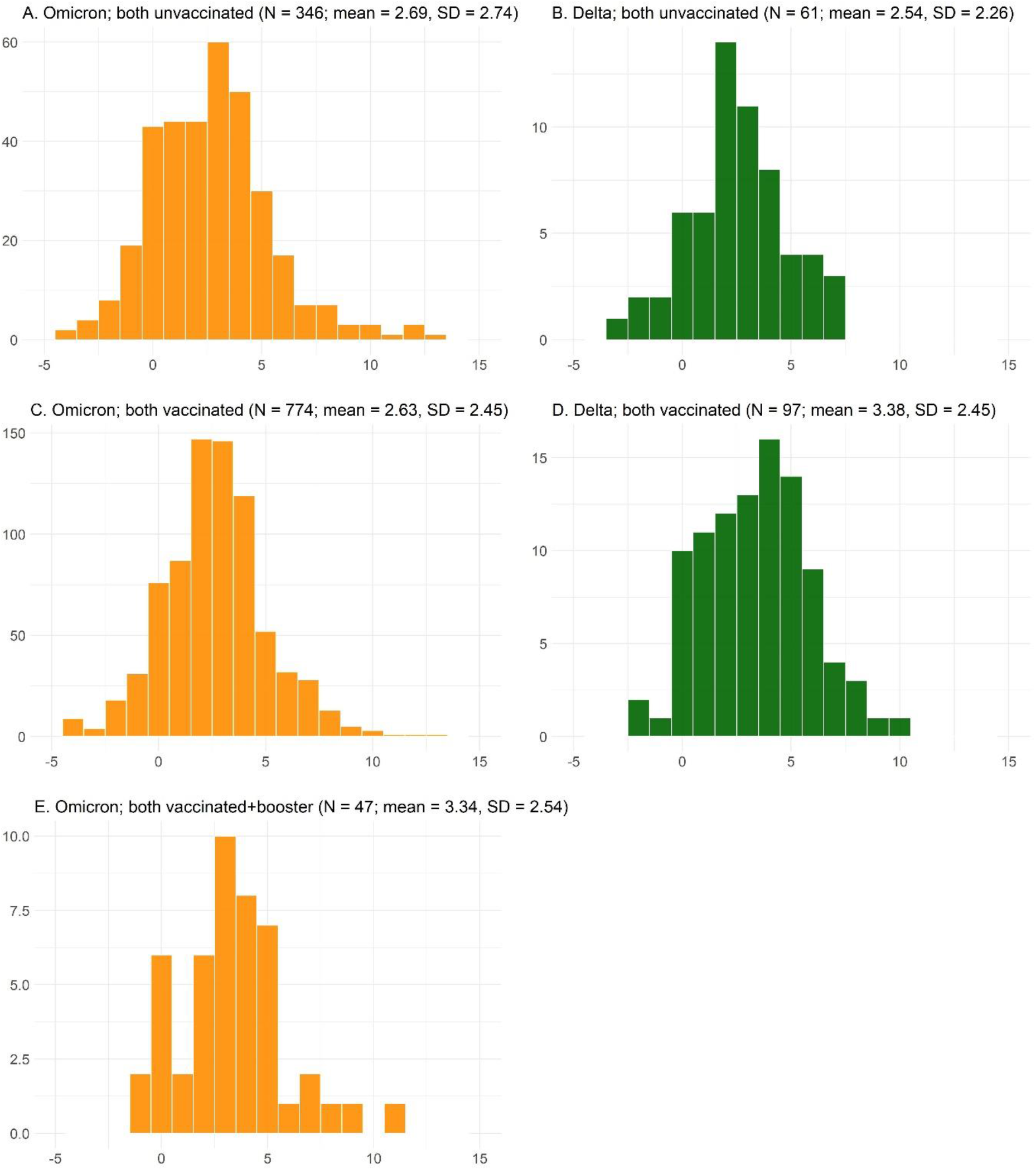
Empirical distribution of the serial intervals for Omicron (A, C, E) and Delta (B, D) variant, for transmission pairs where both cases are unvaccinated (A, B), vaccinated (C, D), or vaccinated with a booster (E, no data for Delta variant). Onset date infector from 19 November until 31 December 2021.

## Discussion

Our estimates of the empirical serial interval for Omicron are in line with those previously reported. Lee et al. [9] reported a mean serial interval of 2.8 days. Based on 18 transmission pairs from South Korea, Kim et al. [10] estimated the mean serial interval to be 2.22 days (95% CrI 1.48 - 2.97) with a standard deviation of 1.62 days (95% CrI 0.87 - 2.37). Backer et al. [7] reported a mean serial interval of 3.5 and 3 days in two consecutive weeks for Omicron within-household pairs. In line with our findings, they found this to be shorter than for Delta pairs. Shorter serial intervals suggest a possibly shorter generation time for the Omicron variant compared to Delta, pointing to faster transmission. This could hence explain the rapid growth that is observed for the Omicron variant. However, control measures and asymptomatic transmission may lead to different serial and generation interval distributions [11]. Future studies estimating the generation time for both variants are needed to shed more light on this.

We found an indication that the empirical serial interval might depend on vaccination status. Precisely, we observed a shorter empirical serial interval for Omicron (2.65 days) compared to Delta (3.38 days) in transmission pairs where both cases completed the vaccination cycle. In addition we found that for the Delta variant the empirical serial interval was longer in transmission pairs where both cases completed the vaccination cycle (3.38 days), compared to those who are unvaccinated or only partially vaccinated (2.54 days). We also found a longer empirical serial interval for Omicron in pairs where both cases already received a booster vaccine (3.34 days) compared to those pairs that were vaccinated but had not yet received a booster (2.63 days). These results indicate that the empirical mean serial interval increases when the infector and infectee have a higher level of vaccine-induced immunity. However, possibly because of limited sample size, this is not observed for all possible combinations of vaccination status (Figure S2) and more data are needed to properly assess the relation between vaccination and serial interval. Interestingly, the empirical mean serial interval for unvaccinated and vaccinated (without booster) transmission pairs was similar for the Omicron variant. If vaccine-induced immunity and serial interval are positively correlated, this result might be explained by lower vaccine efficacy against Omicron for individuals that have not yet received a booster vaccine. While the vaccination campaign progresses and more and more individuals receive a booster vaccine, the reasons for and extent of Omicron’s dominance over Delta might have to be reassessed once more data are available.

This work has several limitations. Date of symptom onset is self-reported by the case and may be subject to recall bias. Likewise, there might be bias in the reporting of contacts and the level of reporting may differ for each index case. We have used all reported transmission pairs, although some of them may have been wrongly assigned. We further assume that directionality of transmission was from index to contact. We also do not explicitly account for right truncation [12], but this is assumed not to affect our estimates since it is unlikely that symptomatic infectees were missed as we used the data available on 17 January 2022, but only including infectors that had symptom onset until 31 December 2021 and limiting the serial interval to at most 15 days. However, because contacts are reconstructed until two days prior to symptom onset of the index case, possible left truncation might lead to exclusion of some transmission pairs. While the transmission risk is higher around symptom onset [8], it is possible that not all the infection events caused by an index case are detected. There is also possible selection bias due to the targeted genotype sequencing of suspected Omicron cases (such as previously infected cases or travelers), whereas genotype sequencing resulting in confirmed Delta cases might have been done on samples from a hospital setting as severe disease was an indication for sequencing during the study period. This analysis does not correct for age and previous infection and hence reinfections might be over-represented among the Omicron cases.

## Data Availability

Transmission pair data (without vaccination status for identifiability reasons) and R code for the analysis are available on https://github.com/cecilekremer/serial_interval

## Ethical and privacy statement

The data analyses carried out in this work are among the legal tasks of Sciensano (artikel 4, §4 Wet tot oprichting van Sciensano; article 4, §4 Law establishment Sciensano). Article 4, §4 explicitly states that Sciensano is authorized to collect, validate, analyse, report and archive data of a personal nature concerning public health. Sciensano is further authorized to make these processed data available with approval of the qualified sectoral committees. Such approval for these data was obtained from the Information Security Committee Social Security and Health (ISC).

## Acknowledgements

Data is provided by Sciensano, the Belgian institute for health. We thank Nicola Low for the insightful comments on an earlier draft of this manuscript.

## Contributions

Study conceptualisation: NH, AT, CK, TB; Literature research: CK, AT, NH; Data collection: TB, KP; Data analysis code: CK; Data analysis: CK, AT, NH; Results interpretation: AT, CK, NH, EA, TB, KP; Manuscript writing: CK, AT, NH; Manuscript review: TB, KP, EA, NH, AT, CK; Coordination: NH. CK and TB contributed equally to this work.

