## Supplementary Material for "Observed serial intervals of SARS-CoV-2 for the Omicron and Delta variants in Belgium based on contact tracing data, 19 November to 31 December 2021"

[†]These authors contributed equally to this work.

### Supplement S1: Detailed description of the data and analysis

During the study period, contacts that were considered high-risk (typically close contact (<1.5m) for longer time periods (>15 min)), were required to take a PCR-test. A first test was taken as soon as possible, a second test, when the first test was negative, was taken seven days after the last contact. In this way, contacts that test positive for SARS-CoV-2 can be linked to a previously confirmed case and transmission pairs can be reconstructed. Vaccination was deemed to be effective after 7 days for Pfizer (second dose), 14 days for Moderna or AstraZeneca (second dose), 21 days for JJ (only one dose), or 7 days for a booster vaccine (Pfizer or Moderna). Other possible combinations seen as a complete vaccination schedule were one dose AstraZeneca followed by a Pfizer dose, or one dose Pfizer followed by Moderna.

We first analyze all transmission pairs for both the Omicron and the Delta variant, reporting the empirical distribution and computing the empirical mean and standard deviation of the observed serial intervals. Furthermore, we fit a normal distribution to the observed serial intervals, accounting for negative serial intervals, i.e. when the infectee shows symptoms before the infector [1]. Normal distributions were fit using Markov

Chain Monte Carlo (MCMC) methods and we report the posterior median and 95% credibility interval (CrI) for the mean and standard deviation. We then compared observed serial intervals for transmission that took place within and between households, where the latter refers to infection between individuals that do not live together. Furthermore, we compared observed serial intervals based on the vaccination status of cases. The significance of differences in observed serial intervals was tested using a Mann-Whitney U test, since the diagnostics for a $t$-test were not met.

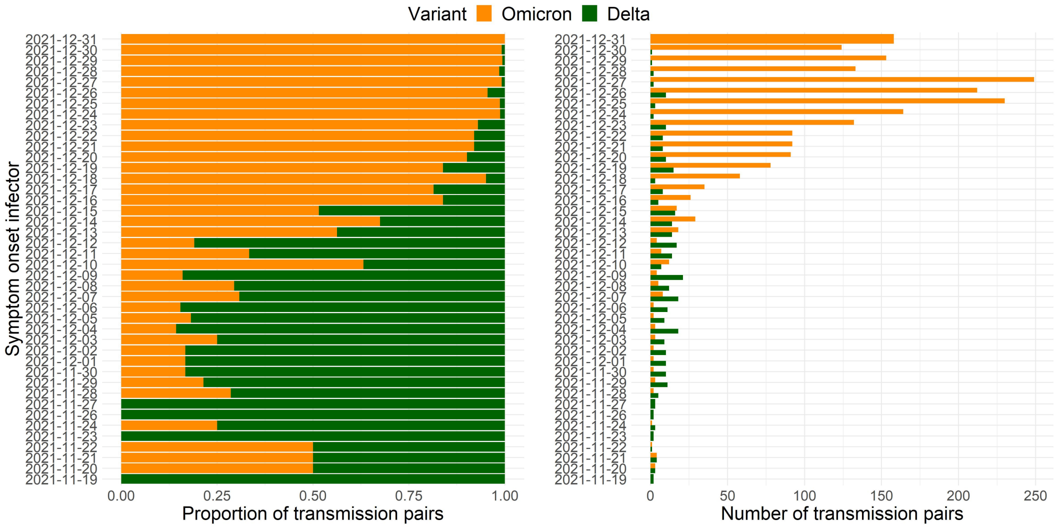


Figure S1: Proportion of all included (left) and total number (right) of transmission pairs linked to Omicron and Delta variant, by infector symptom onset date from 19 November to 31 December 2021.

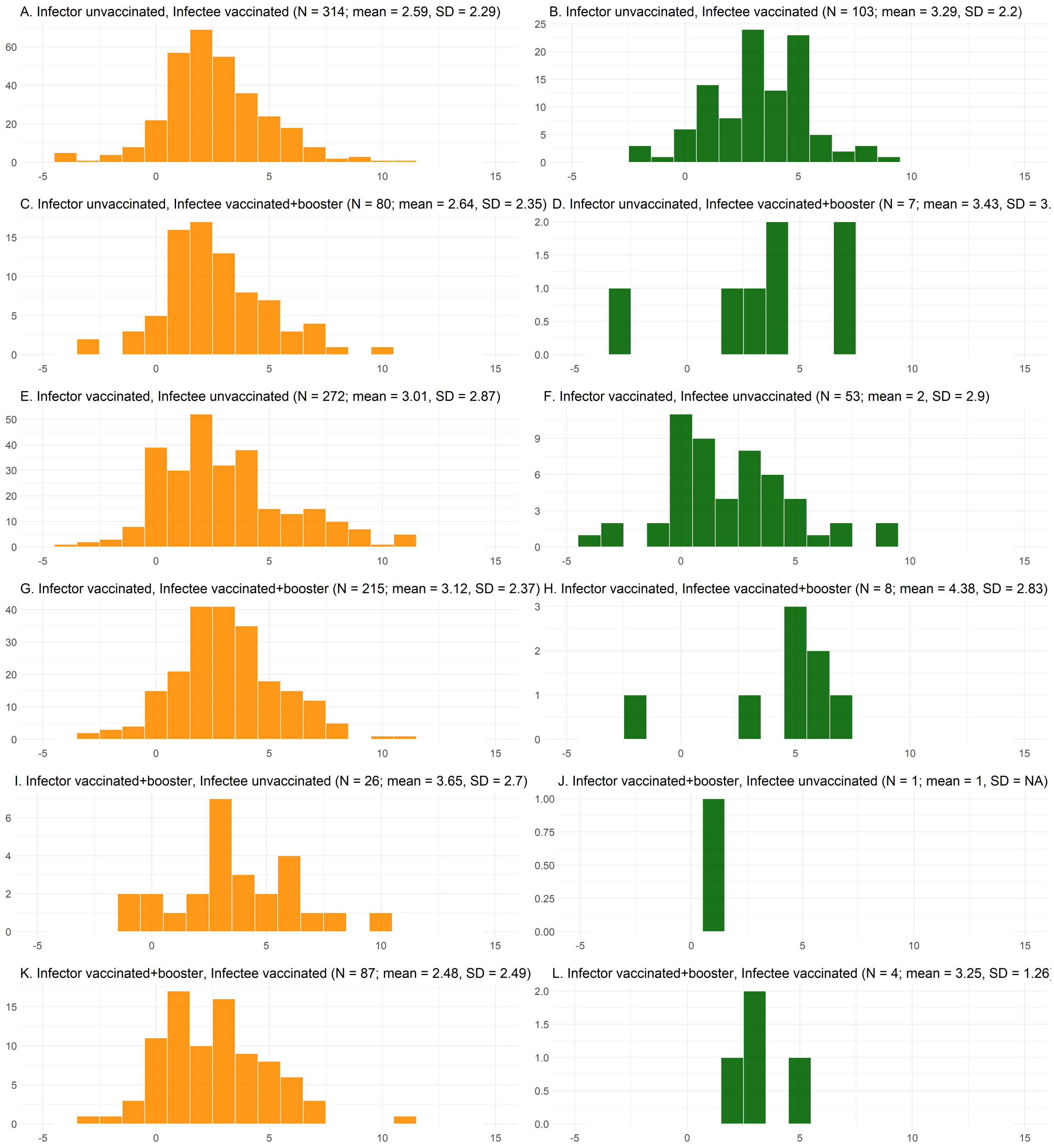


Figure S2: Empirical distribution of the serial intervals for Omicron (A, C, E, G, I, K) and Delta (B, D, F, H, J, L) variant, by vaccination status of infector and infectee. Infector symptom onset from 19 November until 31 December 2021.

Table S1: Proportion of within- and between-household transmission pairs for both variants.

| **Household status** | **Variant** | **Proportion of all pairs** |
|---|---|---|
| Within-household | Omicron | 0.68 |
| | Delta | 0.85 |
| Between-household | Omicron | 0.32 |
| | Delta | 0.15 |

Table S2: Proportion of transmission pairs by vaccination status for both variants.

| **Vaccination status** | **Variant** | **Proportion of all pairs** |
|---|---|---|
| Both unvaccinated | Omicron | 0.16 |
| | Delta | 0.18 |
| Both vaccinated without booster | Omicron | 0.36 |
| | Delta | 0.29 |
| Both vaccinated plus booster | Omicron | 0.02 |
| | Delta | 0.00 |
